# A Lasting Legacy: Long-Term Effects of Exercise Training on Cardiometabolic Health in the STRRIDE-Prediabetes Reunion Study

**DOI:** 10.64898/2026.05.26.26352907

**Authors:** Leanna M. Ross, Alyssa M. Sudnick, Katherine A. Collins-Bennett, Nannan Bo, Julie D. Counts, Johanna L. Johnson, William C. Bennett, Agustin A. Saldana, Katie G. Kennedy, Constantin F. Aliferis, Sisi Ma, Kim M. Huffman, Sarah B. Peskoe, William E. Kraus

**Author notes:** **Corresponding Author**: Leanna M. Ross, PhD, Assistant Professor in Medicine, Division of Cardiology – Department of Medicine, Duke University School of Medicine, 3475 Erwin Road, Aesthetics Bldg., Rm 254, Durham, NC 27705.

## Abstract

**Background:** Regular exercise is a highly effective yet underutilized strategy to reduce cardiometabolic disease burden. Whether brief structured exercise programs confer lasting cardiometabolic benefits remains unclear. The STRRIDE-Prediabetes Reunion study examined legacy effects of exercise training on cardiorespiratory fitness, body composition, and cardiometabolic health.

**Methods:** Seventy-three participants (71.3±7.2 years; 64% women; 77% White) completed Reunion assessments ∼11 years after completing one of four 6-month interventions differing in exercise amount, intensity, and inclusion of diet-induced weight loss. Linear mixed effects models evaluated longitudinal trajectories; secondary analyses examined baseline-adjusted associations among short-term intervention response and Reunion outcomes.

**Results:** Abdominal adiposity improved across all groups from baseline to Reunion, with waist circumference decreasing ∼3 cm over the follow-up period. In contrast, cardiorespiratory fitness and fat-free mass declined significantly. A significant group by time interaction was observed for total fat mass (*p*=0.01), with continued fat mass reductions observed in women randomized to high amount exercise. After baseline adjustment, greater short-term intervention response was associated with more favorable Reunion outcomes across fitness, body composition, and cardiometabolic domains; fat-free mass showed the strongest association (β=0.84, *p*<0.0001).

**Conclusions:** In older adults with prediabetes, the STRRIDE-Prediabetes interventions produced several legacy health effects persisting more than a decade later. Legacy effects differed by sex and exercise dose, and short-term intervention response relative to baseline was associated with long-term outcomes – supporting targeted exercise strategies to preserve cardiometabolic health and functional independence with aging.

## INTRODUCTION

Nearly half of US adults have cardiovascular disease, which remains the leading cause of morbidity and mortality.^1^ Physical inactivity is a key modifiable contributor, responsible for more than 7% of all-cause and cardiovascular disease deaths worldwide.^2^ Although exercise training elicits well-established health benefits,^3^ adoption and maintenance remain critically low – only 26% of men and 19% of women meet current aerobic and resistance exercise guidelines.^4^ Of particular clinical relevance is whether a relatively brief period of structured exercise training, such as a 6-8 month program, can confer persistent health benefits, regardless of long-term physical activity maintenance.

The sustained benefit of a treatment long after cessation is termed a legacy effect.^5^ This concept was established in pharmaceutical trials, including ten-year follow-up of the UK Prospective Diabetes Study – where intensive glucose control produced persistent cardiovascular benefits^6^ – and long-term follow-up of statin trials demonstrating reductions in cardiovascular events and all-cause mortality.^7,8^ Legacy effects have also been observed following intensive lifestyle interventions: the Finnish Diabetes Prevention Study, the Da Qing Diabetes Prevention Study, and the Diabetes Prevention Program (DPP) each reported persistent cardiometabolic benefits a decade or more after trial cessation.^9–12^ Importantly, because these landmark trials combined dietary modification, weight loss goals, and exercise, the independent contribution of exercise training to any observed legacy effects remains unclear.

To date, STRRIDE I (Studies of a Targeted Risk Reduction Intervention Through Defined Exercise) represents the most rigorously characterized example of exercise-induced legacy effects from a randomized controlled trial, with some effects varying by exercise dose and intensity.^13,14^ Participants in vigorous intensity exercise groups experienced only half the expected age-related cardiorespiratory fitness decline (∼4.7% vs. expected ∼10% per decade^15–17^), while moderate intensity exercise yielded the most consistent legacy benefits for resting blood pressure and fasting insulin.^13^ However, exercise-induced legacy effects have yet to be validated in an independent cohort, and whether such effects extend to populations at heightened cardiometabolic risk (*e.g.,* those with prediabetes).

The STRRIDE-Prediabetes (STRRIDE-PD) Reunion study is uniquely positioned to address these questions. The original STRRIDE-PD trial included a combined exercise plus diet-induced weight loss arm designed to mirror the lifestyle intervention of the DPP – the established gold standard for diabetes prevention – alongside three aerobic exercise-only groups differing in amount and intensity.^18^ This design directly evaluates how much of the gold standard lifestyle effect can be achieved through the exercise component alone, and with approximately ten-year follow-up, whether those effects persist. Critically, this design allows legacy effects of exercise training to be examined both independently and in the context of combined lifestyle intervention – complementing and extending the long-term findings of prior diabetes prevention trials in a way previous exercise studies have been unable to capture. Thus, the purpose of this study is to determine whether short-term exercise training – with or without diet-induced weight loss – elicits sustained (legacy) effects on cardiorespiratory fitness, body composition, and cardiometabolic risk factors.

## METHODS

### Study participants

The parent STRRIDE-PD trial (1R01DK081559; NCT00962962; 2009-2014) evaluated the effects of different amounts and intensities of aerobic exercise – with and without diet-induced weight loss – on cardiometabolic health outcomes in sedentary adults (45-75 years) with overweight or obesity and prediabetes (defined by two consecutive fasting glucose concentrations ≥95 to <126 mg/dL obtained one week apart).^18^ The STRRIDE-PD study design and primary outcomes have been previously reported in detail.^18^ Briefly, participants were randomized into one of four 6-month intervention groups: 1) <u>low amount/moderate intensity exercise (Low/Mod)</u>: 10 kcal exercise expenditure/kg body weight/week (KKW) at 40-55% VϑO_2_ reserve; 2) <u>high amount/moderate intensity</u> <u>exercise (High/Mod)</u>: 16 KKW at 40-55% VϑO_2_ reserve; 3) <u>high amount/vigorous intensity exercise</u> <u>(High/Vig)</u>: 16 KKW at 65-80% VϑO_2_ reserve; or 4) <u>clinical lifestyle intervention (Low/Mod+Diet)</u>: 10 KKW at 40-55% VϑO_2_ reserve plus a calorically restricted diet with a weight loss goal. Modeled after the DPP,^19^ the clinical lifestyle intervention targeted 7% body weight loss through energy-intake restriction, a low-fat diet, and exercise. Participants in this group attended four initial counseling sessions followed by 12 intensive group sessions adapted from the DPP intervention manual. For all groups, exercise intensity and duration were verified by direct supervision and/or downloadable heart rate monitors (Polar Electro, Woodbury, NY, USA). Exercise was performed predominantly on treadmills, with elliptical trainers, rowing ergometers, and cycle ergometers used as alternatives.

All participants having completed the STRRIDE-PD trial (n=175; 61.7% women and 17.1% African American) were considered for participation in the STRRIDE-PD Reunion follow-up study (R21AG075379; 2019-2024). Participants were identified using parent trial records and electronic medical record review. Approximately 10 years after original study completion, participants were contacted via recruitment letters, with follow-up calls providing a study overview. Interested participants were scheduled for an information and consent session. The study protocol was approved by the Duke University Health System Institutional Review Board (Pro00014088). All participants provided written informed consent.

### STRRIDE-PD Reunion Assessments

#### Medical history

Participants completed a 10-year medical history questionnaire, including current medication use.

#### Physical activity

Recent physical activity was assessed using a 23-item questionnaire evaluating frequency, duration, mode, and intensity (expansion of the Paffenbarger Physical Activity Questionnaire^20^). As an objective assessment of current free-living physical activity, participants’ daily step counts were measured and recorded with a wrist-worn Garmin device (vívosmart^®^ 5). Participants were asked to wear the device 24 hours per day (except for brief removal for charging) for at least 7 days between in-person assessment visits. Data retrieval from the device was facilitated by the Garmin Connect and Pattern Health applications (Garmin Connect: https://connect.garmin.com; Pattern Health: https://pattern.health). Raw epoch-level data from the Garmin device were imported, timestamps standardized, and a complete sequence of 15-minute epochs reconstructed for each calendar day. Wear time was inferred from observed step patterns, defined as the interval between the first and last non-zero step epochs within each day. A calendar day was classified as valid if at least 10 hours of inferred wear time were observed, and participants contributing fewer than four valid days were excluded from step-count analyses.^21^

#### Anthropometrics and vital signs

While wearing lightweight clothing and no shoes, height and body weight were measured with a stadiometer and digital scale, respectively. Waist circumference was measured with a flexible tape measure at the minimal waist – defined as the smallest horizontal circumference above the umbilicus and below the xiphoid process.^22^ Blood pressure was assessed after approximately five minutes of seated rest, with two readings obtained five minutes apart; the average of the two readings was used for analysis.

#### Blood-based markers

Phlebotomy was performed following a 12-hour overnight fast. Glycated hemoglobin (HbA1c) was determined from whole blood via turbidimetric immunoassay (Labcorp). Glucose was determined via enzymatic assay with a clinical analyzer (Labcorp). Plasma was immediately isolated via centrifugation and stored at −80°C. Stored plasma samples from both the parent STRRIDE-PD trial and the Reunion study were analyzed for lipids and lipoproteins using nuclear magnetic resonance LipoProfile testing (LP4 algorithm^23^ on a Vantera Clinical Analyzer,^24^ Labcorp).

#### Body composition

Fat mass and fat-free mass were assessed by air displacement plethysmography (BOD POD^®^ GS-X, Concord, CA). BOD POD^®^ assessments were incorporated after Reunion study initiation; consequently, body composition data at the Reunion timepoint were unavailable for the first five participants enrolled.

#### Cardiorespiratory fitness

Under medical supervision, participants completed a graded maximal cardiopulmonary exercise test on a treadmill with 12-lead electrocardiography and expired gas analysis (TrueOne 2400 Parvomedics; Provo, UT). The graded testing protocol began at 3 mph and 0% grade, with 2-minute stages incrementally increasing speed and/or grade by approximately one metabolic equivalent per stage until volitional exhaustion. If warranted – due to participant health status and/or mobility concerns – an alternate graded protocol with a slower starting speed was administered. Peak VϑO_2_ was determined as the average of the two highest consecutive 15-second values in the last 90-seconds of the test.^25^

#### Metabolic syndrome

Cardiometabolic disease risk was evaluated using a composite metabolic syndrome *z*-score (MSSc) – a continuous weighted score incorporating five metabolic syndrome components: waist circumference, mean arterial blood pressure, fasting blood glucose, triglycerides (TG), and high-density lipoprotein cholesterol (HDL-C).^26^ Sex-specific modified *z*-scores were calculated using the continuous differences between each participant’s values and the Adult Treatment Panel III guideline threshold, normalized to the baseline standard deviation (SD) derived from STRRIDE-PD participants who later completed the Reunion study. A negative score indicates lower cardiometabolic risk.

Women’s MSSc = [(50 – HDL-C)/11.9] + [(TG −150)/53.7] + [(fasting blood glucose − 100)/8.3] + [(waist circumference − 88)/6.9] + [(mean arterial pressure – 100)/9.7]

Men’s MSSc = [(40 – HDL-C)/7.3] + [(TG −150)/53.7] + [(fasting blood glucose − 100)/8.3] + [(waist circumference − 102)/7.7] + [(mean arterial pressure − 100)/9.7]

#### Statistical Analysis

Outcomes of interest spanned four domains: body composition and anthropometrics [percent fat mass, percent fat-free mass, total fat mass, total fat-free mass, waist circumference, and body mass index (BMI)]; cardiorespiratory fitness (absolute peak VϑO₂ in L/min and relative peak VϑO₂ in mL/kg/min); cardiometabolic blood-based markers [TG, total cholesterol, low-density lipoprotein cholesterol (LDL-C), HDL-C, and fasting glucose]; and hemodynamic measures (resting mean arterial pressure, systolic blood pressure, and diastolic blood pressure). Analyses were exploratory in nature, with particular interest in whether trajectories varied by intervention group and sex over time. Accordingly, no formal sample size or power calculations were performed.

Participant characteristics at the time of the original STRRIDE-PD trial baseline and at the Reunion timepoint are presented as means ± SD for continuous variables and frequencies with percentages for categorical variables. Differences in baseline characteristics across intervention groups were assessed using Kruskal-Wallis tests for continuous variables and Chi-square tests for categorical variables.

Linear mixed effects models were fit separately for each outcome, with random effects for participant and fixed effects for intervention group, sex, and time, as well as interactions between intervention group and time, and between sex and time. The following effects were tested using partial F-tests, with statistical significance defined as *p* < 0.05: main effects of time, intervention group, and sex; the intervention group by time interaction; and the sex by time interaction. All models were estimated using maximum likelihood with an unstructured covariance matrix. To account for potential confounding by pharmacotherapy, models for blood pressure, lipid, and glucose outcomes were adjusted for relevant medication use (yes/no) at the Reunion timepoint. Given the exploratory nature of these analyses, no correction for multiple comparisons was applied and acknowledgements of potential associations are noted for (0.05 ≤ *p* < 0.10) alongside significant findings (*p* < 0.05 for main effects, and *p* < 0.10 for interaction effects).

To examine the association between the original intervention response and long-term outcomes, a separate linear regression model was fit for each outcome across all participants, adjusting for the corresponding baseline value from the original STRRIDE-PD trial. Specifically, the model took the form: Y_reunion_i_ = β₀ + β₁·ChangeL + β₂·Y_baseline_i_ + εL, where β₁ represents the mean difference in the Reunion outcome per one-unit increase in the baseline-to-post-intervention change, holding baseline value constant, and εL is the residual error term.

To facilitate direct comparison with previously reported legacy effects on cardiorespiratory fitness from the STRRIDE I Reunion cohort,^13^ percent change in relative peak VϑO_2_ from baseline to the Reunion timepoint was calculated for each participant as [(Reunion value − baseline value) / baseline value] × 100 and summarized descriptively by intervention group. One-sample t-tests were used to evaluate whether mean percent change within each group was significantly different from zero. This analysis was descriptive in nature and not subject to correction for multiple comparisons.

Analyses were conducted as complete cases, restricted to participants who completed the Reunion study, and a descriptive comparison between those who did and did not complete the Reunion study was performed. Outcome-specific observed sample sizes among those who completed the Reunion study were reported. All analyses were performed using SAS statistical software (SAS Institute, Cary, NC).

## RESULTS

Of the 175 participants who completed the original STRRIDE-PD trial, 4 were deceased, 11 had relocated out of state, 5 were unable to participate due to health reasons, and 70 were unable to be contacted or did not respond. Of the remaining 85 eligible participants, 7 declined, 3 were lost to follow-up, and 75 provided informed consent. Subsequently, 2 participants withdrew – one due to health reasons and one for personal reasons – leaving 73 participants who completed Reunion assessments. Of these, 6 did not complete the cardiopulmonary exercise test: 3 elected not to participate in this assessment and 3 were unable to complete testing due to mobility limitations or claustrophobia. On average, participants returned for Reunion assessments 11.4 ± 1.7 years after completing the original STRRIDE-PD trial. Descriptive comparison of baseline characteristics and intervention response measures between the 73 Reunion participants and the 175 original STRRIDE-PD completers revealed no meaningful differences, supporting the representativeness of the Reunion cohort (**Table S1**).

The 73 Reunion participants (71.3 ± 7.2 years; 76.7% White) included 47 women and 26 men. The most commonly reported medical conditions at the Reunion timepoint were abnormal cholesterol (47.9%), hypertension (45.2%), cancer (19.2%), and high TG (13.7%). Notably, only 8 of 73 participants (11.0%) reported having a diagnosis of type 2 diabetes at the Reunion timepoint, including 3 of 18 participants in the High/Mod group (16.7%), 2 of 23 in the High/Vig group (8.7%), 0 of 15 in the Low/Mod group (0%), and 3 of 17 in the Low/Mod+Diet group (17.6%). Overall, 72.6% of participants reported current use of at least one cardiometabolic medication. Specific medication classes included blood pressure-lowering medications (52.1%), lipid-lowering medications (45.2%), and glucose-lowering medications (11.0%). At the Reunion timepoint, participants self-reported physical activity frequency in the past 3 months as follows: 17.8% exercised 0-1 days/week, 49.3% exercised 2-3 days/week, and 32.9% exercised 4 or more days/week. On average, participants accumulated 5,964 ± 2,867 steps per day across a median of 12 valid wear days, with considerable individual variability; average daily step counts did not differ meaningfully across intervention groups. **Table 1** displays participant characteristics at the time of the Reunion study for the total sample and by intervention group.

**Table 1.**
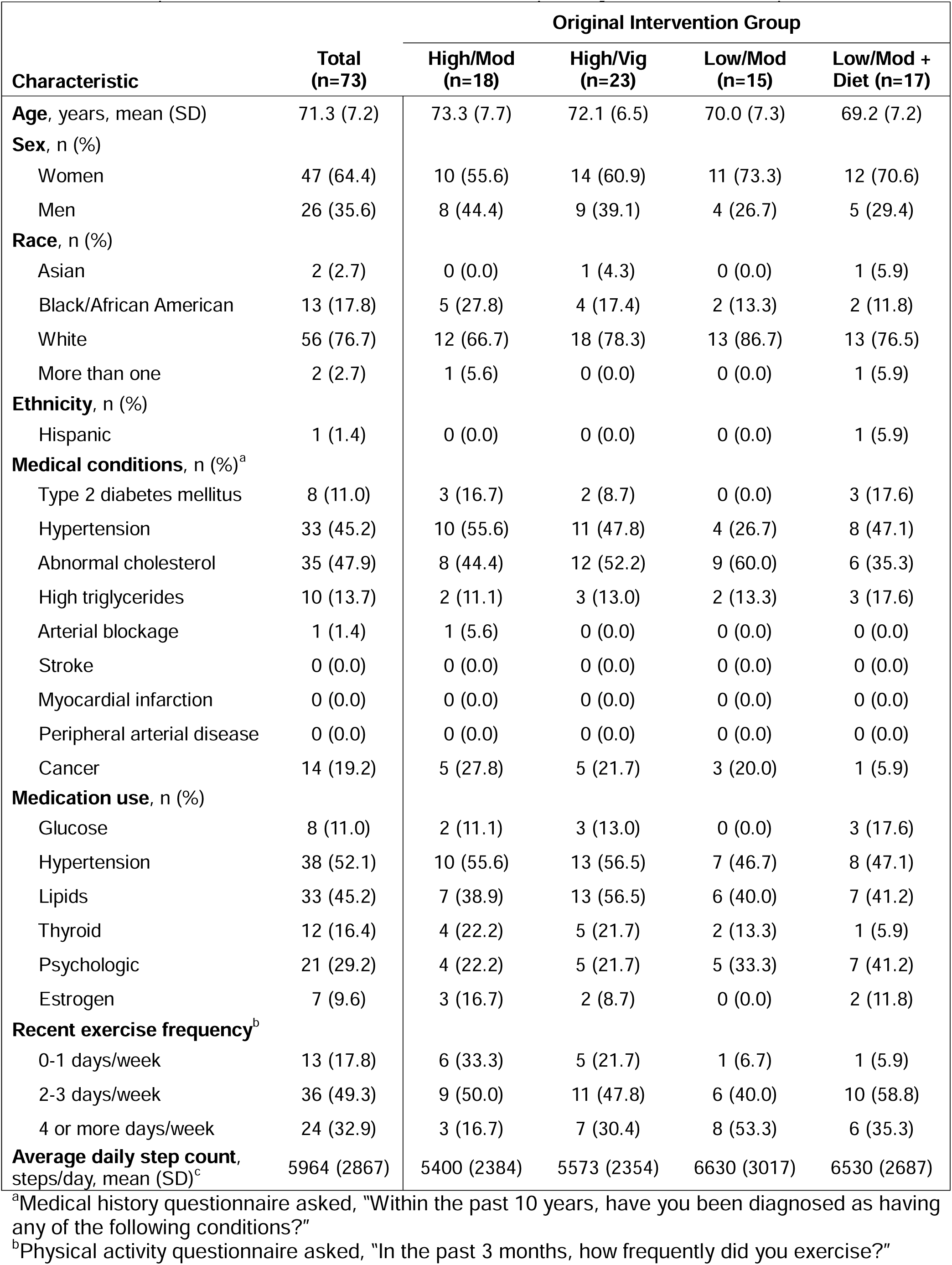
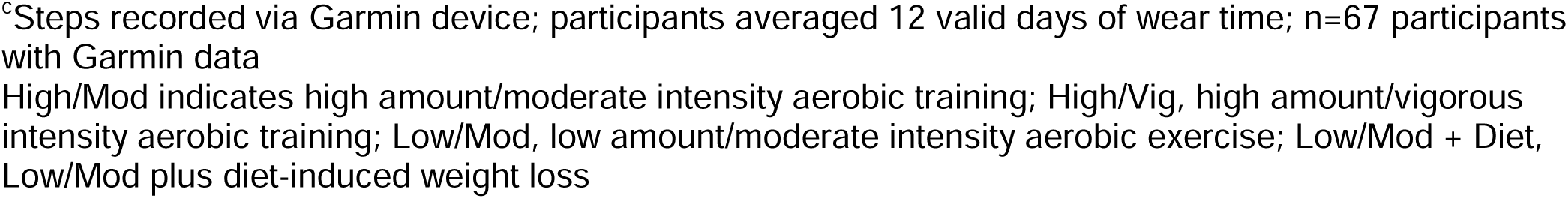
Participant Characteristics at the Reunion Timepoint by Intervention Group.

| Characteristic | Total<br>(n=73) | Original Intervention Group |  |  |  |
| --- | --- | --- | --- | --- | --- |
|  |  | High/Mod<br>(n=18) | High/Vig<br>(n=23) | Low/Mod<br>(n=15) | Low/Mod +<br>Diet (n=17) |
| **Age**, years, mean (SD) | 71.3 (7.2) | 73.3 (7.7) | 72.1 (6.5) | 70.0 (7.3) | 69.2 (7.2) |
| **Sex**, n (%) |  |  |  |  |  |
| Women | 47 (64.4) | 10 (55.6) | 14 (60.9) | 11 (73.3) | 12 (70.6) |
| Men | 26 (35.6) | 8 (44.4) | 9 (39.1) | 4 (26.7) | 5 (29.4) |
| **Race**, n (%) |  |  |  |  |  |
| Asian | 2 (2.7) | 0 (0.0) | 1 (4.3) | 0 (0.0) | 1 (5.9) |
| Black/African American | 13 (17.8) | 5 (27.8) | 4 (17.4) | 2 (13.3) | 2 (11.8) |
| White | 56 (76.7) | 12 (66.7) | 18 (78.3) | 13 (86.7) | 13 (76.5) |
| More than one | 2 (2.7) | 1 (5.6) | 0 (0.0) | 0 (0.0) | 1 (5.9) |
| **Ethnicity**, n (%) |  |  |  |  |  |
| Hispanic | 1 (1.4) | 0 (0.0) | 0 (0.0) | 0 (0.0) | 1 (5.9) |
| **Medical conditions**, n (%)<sup>a</sup> |  |  |  |  |  |
| Type 2 diabetes mellitus | 8 (11.0) | 3 (16.7) | 2 (8.7) | 0 (0.0) | 3 (17.6) |
| Hypertension | 33 (45.2) | 10 (55.6) | 11 (47.8) | 4 (26.7) | 8 (47.1) |
| Abnormal cholesterol | 35 (47.9) | 8 (44.4) | 12 (52.2) | 9 (60.0) | 6 (35.3) |
| High triglycerides | 10 (13.7) | 2 (11.1) | 3 (13.0) | 2 (13.3) | 3 (17.6) |
| Arterial blockage | 1 (1.4) | 1 (5.6) | 0 (0.0) | 0 (0.0) | 0 (0.0) |
| Stroke | 0 (0.0) | 0 (0.0) | 0 (0.0) | 0 (0.0) | 0 (0.0) |
| Myocardial infarction | 0 (0.0) | 0 (0.0) | 0 (0.0) | 0 (0.0) | 0 (0.0) |
| Peripheral arterial disease | 0 (0.0) | 0 (0.0) | 0 (0.0) | 0 (0.0) | 0 (0.0) |
| Cancer | 14 (19.2) | 5 (27.8) | 5 (21.7) | 3 (20.0) | 1 (5.9) |
| **Medication use**, n (%) |  |  |  |  |  |
| Glucose | 8 (11.0) | 2 (11.1) | 3 (13.0) | 0 (0.0) | 3 (17.6) |
| Hypertension | 38 (52.1) | 10 (55.6) | 13 (56.5) | 7 (46.7) | 8 (47.1) |
| Lipids | 33 (45.2) | 7 (38.9) | 13 (56.5) | 6 (40.0) | 7 (41.2) |
| Thyroid | 12 (16.4) | 4 (22.2) | 5 (21.7) | 2 (13.3) | 1 (5.9) |
| Psychologic | 21 (29.2) | 4 (22.2) | 5 (21.7) | 5 (33.3) | 7 (41.2) |
| Estrogen | 7 (9.6) | 3 (16.7) | 2 (8.7) | 0 (0.0) | 2 (11.8) |
| **Recent exercise frequency**<sup>b</sup> |  |  |  |  |  |
| 0-1 days/week | 13 (17.8) | 6 (33.3) | 5 (21.7) | 1 (6.7) | 1 (5.9) |
| 2-3 days/week | 36 (49.3) | 9 (50.0) | 11 (47.8) | 6 (40.0) | 10 (58.8) |
| 4 or more days/week | 24 (32.9) | 3 (16.7) | 7 (30.4) | 8 (53.3) | 6 (35.3) |
| **Average daily step count**,<br>steps/day, mean (SD)<sup>c</sup> | 5964 (2867) | 5400 (2384) | 5573 (2354) | 6630 (3017) | 6530 (2687) |

<sup>a</sup>Medical history questionnaire asked, "Within the past 10 years, have you been diagnosed as having any of the following conditions?"
<sup>b</sup>Physical activity questionnaire asked, "In the past 3 months, how frequently did you exercise?"
<sup>c</sup>Steps recorded via Garmin device; participants averaged 12 valid days of wear time; n=67 participants with Garmin data
High/Mod indicates high amount/moderate intensity aerobic training; High/Vig, high amount/vigorous intensity aerobic training; Low/Mod, low amount/moderate intensity aerobic exercise; Low/Mod + Diet, Low/Mod plus diet-induced weight loss

### Longitudinal Outcomes

Outcome values at baseline, post-intervention, and Reunion are presented for the total sample in **Table 2a** and by intervention group in **Table 2b**; *p*-values from linear mixed effects models are presented in **Table 3**. Outcome-specific observed sample sizes are included both overall (**Table 2a**) and by intervention group (**Table 2b**).

**Table 2a.** Longitudinal Outcome Measures at Baseline, Post-Intervention, and Reunion.

| **Outcome** | **Baseline** | **Post** | **Reunion** | **n** |
| --- | --- | --- | --- | --- |
| Body Weight (kg) | 86.0 (12.9) | 83.3 (12.0) | 82.2 (14.4) | 73 |
| BMI (kg/m<sup>2</sup>) | 30.2 (2.8) | 29.2 (2.8) | 29.1 (3.4) | 73 |
| Minimal Waist Circumference (cm) | 98.2 (9.4) | 96.3 (8.9) | 95.0 (11.1) | 68 |
| Fat Mass (%) | 41.2 (7.6) | 39.5 (7.9) | 41.5 (7.5) | 66 |
| Fat-free Mass (%) | 58.8 (7.6) | 60.5 (7.9) | 58.5 (7.5) | 66 |
| Total Fat Mass (kg) | 35.4 (7.3) | 32.8 (7.2) | 33.8 (8.3) | 66 |
| Total Fat-free Mass (kg) | 51.4 (11.3) | 50.7 (11.1) | 47.8 (11.2) | 66 |
| Absolute Peak $\dot{V}\text{O}_2$ (L/min) | 2.16 (0.65) | 2.32 (0.67) | 1.81 (0.59) | 73 |
| Relative Peak $\dot{V}\text{O}_2$ (mL/kg/min) | 24.7 (5.2) | 27.4 (5.6) | 21.9 (5.4) | 73 |
| Metabolic Syndrome z-score | -0.2 (2.8) | -1.4 (3.0) | -1.7 (4.1) | 64 |
| Fasting Glucose (mg/dL) | 104.4 (8.3) | 103.8 (9.3) | 105.6 (16.3) | 72 |
| HDL-C (mg/dL) | 47.7 (11.0) | 49.6 (11.3) | 62.7 (19.3) | 72 |
| LDL-C (mg/dL) | 98.7 (21.6) | 98.1 (18.8) | 98.7 (31.7) | 72 |
| Total Cholesterol (mg/dL) | 173.0 (26.6) | 173.2 (24.2) | 186.8 (38.8) | 72 |
| Total Triglycerides (mg/dL) | 119.2 (53.7) | 109.5 (55.1) | 126.9 (85.1) | 72 |
| Resting MAP (mmHg) | 91.2 (9.7) | 89.0 (9.4) | 95.2 (9.3) | 68 |
| Resting Systolic BP (mmHg) | 123.6 (12.7) | 121.4 (14.3) | 130.2 (14.7) | 68 |
| Resting Diastolic BP (mmHg) | 74.9 (9.8) | 73.1 (8.8) | 77.7 (9.3) | 68 |

For each outcome, results are presented for *n* participants with data available at all three timepoints; values presented as mean (standard deviation); BMI indicates body mass index; BP, blood pressure; HDL-C, high-density lipoprotein cholesterol; LDL-C, low-density lipoprotein cholesterol; MAP, mean arterial pressure; Post, post-intervention; $\dot{V}\text{O}_2$ , oxygen consumption

**Table 2b.** Longitudinal Outcome Measures at Baseline, Post-Intervention, and Reunion by Intervention Group.

| Outcome | Original Intervention Group |  |  |  |  |  |  |  |  |  |  |  |  |  |  |  |
| --- | --- | --- | --- | --- | --- | --- | --- | --- | --- | --- | --- | --- | --- | --- | --- | --- |
|  | High/Mod |  |  |  | High/Vig |  |  |  | Low/Mod |  |  |  | Low/Mod + Diet |  |  |  |
|  | Baseline | Post | Reunion | n | Baseline | Post | Reunion | n | Baseline | Post | Reunion | n | Baseline | Post | Reunion | n |
| Body Weight (kg) | 88.7<br>(12.6) | 87.2<br>(11.8) | 84.8<br>(17.2) | 18 | 83.9<br>(12.1) | 81.8<br>(12.2) | 79.7<br>(13.9) | 23 | 85.8<br>(11.5) | 84.2<br>(10.4) | 81.9<br>(12.1) | 15 | 86.2<br>(15.7) | 80.4<br>(13.2) | 83.1<br>(14.5) | 17 |
| BMI (kg/m<sup>2</sup>) | 30.9<br>(2.2) | 30.1<br>(2.0) | 29.8<br>(3.7) | 18 | 29.7<br>(3.1) | 29.0<br>(3.4) | 28.5<br>(3.4) | 23 | 30.6<br>(2.6) | 30.0<br>(2.4) | 29.5<br>(2.8) | 15 | 29.9<br>(3.3) | 27.9<br>(2.7) | 29.0<br>(3.6) | 17 |
| Minimal Waist Circumference (cm) | 99.7<br>(8.8) | 97.4<br>(9.2) | 95.9<br>(12.1) | 16 | 98.2<br>(9.1) | 96.8<br>(8.8) | 95.2<br>(10.8) | 21 | 99.4<br>(9.3) | 97.7<br>(9.0) | 95.4<br>(10.8) | 15 | 95.4<br>(10.4) | 93.2<br>(8.9) | 93.6<br>(11.5) | 16 |
| Fat Mass (%) | 40.6<br>(6.3) | 39.2<br>(7.0) | 41.3<br>(6.6) | 16 | 41.4<br>(9.2) | 39.1<br>(9.7) | 39.7<br>(8.4) | 22 | 41.9<br>(6.4) | 42.0<br>(6.9) | 44.0<br>(6.7) | 14 | 41.1<br>(7.9) | 38.0<br>(6.4) | 42.1<br>(7.7) | 14 |
| Fat-free Mass (%) | 59.4<br>(6.3) | 60.8<br>(7.0) | 58.7<br>(6.6) | 16 | 58.6<br>(9.2) | 60.9<br>(9.7) | 60.3<br>(8.4) | 22 | 58.1<br>(6.4) | 58.0<br>(6.9) | 56.0<br>(6.7) | 14 | 58.9<br>(7.9) | 62.0<br>(6.4) | 57.9<br>(7.7) | 14 |
| Total Fat Mass (kg) | 36.0<br>(4.5) | 33.7<br>(5.7) | 34.2<br>(7.7) | 16 | 34.4<br>(9.1) | 31.8<br>(9.6) | 31.5<br>(9.7) | 22 | 35.5<br>(5.6) | 34.7<br>(5.9) | 35.4<br>(6.0) | 14 | 36.4<br>(8.8) | 31.3<br>(5.3) | 34.8<br>(8.5) | 14 |
| Total Fat-free Mass (kg) | 54.0<br>(12.1) | 53.3<br>(11.8) | 49.4<br>(13.3) | 16 | 49.0<br>(10.9) | 49.6<br>(11.0) | 47.9<br>(10.7) | 22 | 50.0<br>(11.0) | 48.6<br>(10.4) | 45.8<br>(11.4) | 14 | 53.5<br>(11.3) | 52.0<br>(11.7) | 47.7<br>(10.3) | 14 |
| Absolute Peak V̇O<sub>2</sub> (L/min) | 2.20<br>(0.72) | 2.34<br>(0.62) | 1.74<br>(0.61) | 18 | 2.05<br>(0.58) | 2.34<br>(0.65) | 1.75<br>(0.46) | 23 | 2.14<br>(0.63) | 2.27<br>(0.77) | 1.83<br>(0.68) | 15 | 2.30<br>(0.72) | 2.32<br>(0.73) | 1.96<br>(0.65) | 17 |
| Relative Peak V̇O<sub>2</sub> (mL/kg/min) | 24.2<br>(5.1) | 26.4<br>(4.4) | 20.2<br>(5.2) | 18 | 24.1<br>(4.8) | 27.9<br>(5.6) | 21.9<br>(4.3) | 23 | 24.6<br>(4.9) | 26.4<br>(6.0) | 22.0<br>(6.0) | 15 | 26.3<br>(6.1) | 28.5<br>(6.2) | 23.7<br>(6.0) | 17 |
| Metabolic Syndrome z-score | 0.1<br>(2.2) | -1.5<br>(2.8) | -2.3<br>(3.9) | 15 | -1.0<br>(3.3) | -1.7<br>(3.6) | -2.1<br>(5.0) | 19 | 0.6<br>(2.8) | -0.3<br>(2.9) | -1.1<br>(2.8) | 15 | -0.5<br>(2.3) | -2.1<br>(2.2) | -0.8<br>(4.4) | 15 |
| Fasting Glucose (mg/dL) | 106.3<br>(7.8) | 104.7<br>(9.1) | 105.9<br>(21.0) | 18 | 103.7<br>(9.6) | 103.8<br>(10.4) | 104.7<br>(16.7) | 23 | 105.9<br>(6.2) | 107.6<br>(9.3) | 109.3<br>(11.8) | 15 | 101.7<br>(8.2) | 99.4<br>(6.4) | 103.2<br>(14.2) | 16 |
| HDL-C (mg/dL) | 47.5<br>(11.0) | 49.6<br>(10.8) | 63.9<br>(18.9) | 18 | 50.7<br>(10.7) | 52.5<br>(10.8) | 66.2<br>(21.3) | 23 | 47.2<br>(11.7) | 48.6<br>(12.4) | 61.4<br>(18.0) | 15 | 44.0<br>(10.4) | 46.2<br>(11.6) | 57.7<br>(18.7) | 16 |
| LDL-C (mg/dL) | 101.8<br>(20.6) | 99.4<br>(19.5) | 97.7<br>(33.3) | 18 | 97.6<br>(19.8) | 93.1<br>(15.3) | 98.0<br>(23.5) | 23 | 99.9<br>(23.5) | 100.5<br>(18.5) | 101.1<br>(41.1) | 15 | 95.5<br>(24.7) | 101.2<br>(23.0) | 98.8<br>(33.1) | 16 |
| Total Cholesterol (mg/dL) | 176.9<br>(25.9) | 176.4<br>(24.5) | 184.1<br>(40.6) | 18 | 173.2<br>(24.9) | 170.9<br>(22.3) | 189.8<br>(29.6) | 23 | 174.5<br>(30.5) | 175.0<br>(26.9) | 184.9<br>(50.4) | 15 | 167.1<br>(27.4) | 170.7<br>(25.8) | 187.1<br>(39.8) | 16 |
| Total Triglycerides (mg/dL) | 124.2<br>(66.6) | 117.1<br>(66.1) | 116.1<br>(41.2) | 18 | 111.3<br>(54.1) | 112.0<br>(61.5) | 139.5<br>(129.4) | 23 | 127.6<br>(57.2) | 115.3<br>(50.4) | 108.4<br>(45.8) | 15 | 116.9<br>(33.1) | 90.9<br>(31.2) | 138.1<br>(70.5) | 16 |
| Resting MAP (mmHg) | 89.8<br>(7.5) | 89.8<br>(7.8) | 93.3<br>(4.9) | 15 | 90.8<br>(10.3) | 89.1<br>(9.0) | 96.6<br>(10.2) | 21 | 93.1<br>(7.6) | 88.5<br>(9.4) | 94.3<br>(8.5) | 15 | 91.1<br>(12.4) | 88.5<br>(12.6) | 95.9<br>(12.3) | 17 |
| Resting Systolic BP (mmHg) | 124.1<br>(15.3) | 124.3<br>(13.5) | 132.3<br>(13.4) | 15 | 123.9<br>(12.5) | 124.1<br>(14.0) | 133.1<br>(16.8) | 21 | 121.4<br>(13.2) | 122.0<br>(14.9) | 130.8<br>(16.6) | 15 | 121.8<br>(14.7) | 118.1<br>(17.8) | 127.5<br>(16.4) | 17 |
| Resting Diastolic BP (mmHg) | 72.7<br>(6.6) | 72.5<br>(7.6) | 73.9<br>(4.7) | 15 | 74.2<br>(10.8) | 72.3<br>(8.1) | 78.4<br>(8.7) | 21 | 77.1<br>(8.7) | 74.4<br>(9.5) | 78.5<br>(10.8) | 15 | 75.8<br>(11.8) | 73.6<br>(11.2) | 80.1<br>(11.6) | 17 |

For each outcome, results are presented for $n$ participants with data available at all three timepoints; values presented as mean (standard deviation); BMI indicates body mass index; BP, blood pressure; HDL-C, high-density lipoprotein cholesterol; High/Mod, high amount/moderate intensity aerobic training; High/Vig, high amount/vigorous intensity aerobic training; LDL-C, low-density lipoprotein cholesterol; Low/Mod, low amount/moderate intensity aerobic exercise; Low/Mod + Diet, Low/Mod plus diet-induced weight loss; MAP, mean arterial pressure; Post, post-intervention; $\dot{V}\text{O}_2$ , oxygen consumption

**Table 3.** *P*-values from Linear Mixed Effects Models for Outcome Measures Over Time by Intervention Group and Sex.

| Outcome | P-Value |  |  |  |  |
| --- | --- | --- | --- | --- | --- |
|  | Time (Visit) | Intervention Group | Intervention Group*Time | Sex | Sex*Time |
| Body Weight (kg)<sup>G</sup> | < 0.0001 | 0.38 | < 0.0001 | < 0.0001 | 0.01 |
| BMI (kg/m<sup>2</sup>)<sup>G</sup> | < 0.0001 | 0.33 | 0.0002 | 0.17 | 0.07 |
| Minimal Waist Circumference (cm)<sup>G</sup> | < 0.0001 | 0.48 | 0.19 | < 0.0001 | 0.20 |
| Fat Mass (%)<sup>G</sup> | < 0.0001 | 0.67 | 0.08 | < 0.0001 | 0.06 |
| Fat-free Mass (%)<sup>G</sup> | < 0.0001 | 0.67 | 0.08 | < 0.0001 | 0.06 |
| Total Fat Mass (kg)<sup>G</sup> | < 0.0001 | 0.68 | 0.01 | 0.22 | 0.007 |
| Total Fat-free Mass (kg)<sup>G</sup> | < 0.0001 | 0.18 | 0.19 | < 0.0001 | 0.77 |
| Absolute Peak V̇O<sub>2</sub> (L/min) | < 0.0001 | 0.06 | 0.02 | < 0.0001 | 0.002 |
| Relative Peak V̇O<sub>2</sub> (mL/kg/min) | < 0.0001 | 0.04 | 0.18 | < 0.0001 | 0.05 |
| Metabolic Syndrome z-score<sup>HG</sup> | < 0.0001 | 0.27 | 0.10 | 0.32 | 0.05 |
| Fasting Glucose (mg/dL)<sup>G</sup> | 0.10 | 0.11 | 0.27 | 0.21 | 0.003 |
| HDL-C (mg/dL) | < 0.0001 | 0.16 | 0.99 | 0.003 | 0.23 |
| LDL-C (mg/dL)<sup>L</sup> | 0.97 | 0.97 | 0.37 | 0.001 | 0.78 |
| Total Cholesterol (mg/dL)<sup>L</sup> | 0.03 | 0.79 | 0.88 | 0.001 | 0.995 |
| Total Triglycerides (mg/dL) | 0.001 | 0.997 | 0.09 | 0.03 | 0.05 |
| Resting Mean Arterial Pressure (mmHg)<sup>H</sup> | < 0.0001 | 0.76 | 0.32 | 0.04 | 0.55 |
| Resting Systolic Blood Pressure (mmHg)<sup>H</sup> | 0.0004 | 0.58 | 0.15 | 0.16 | 0.89 |
| Resting Diastolic Blood Pressure (mmHg)<sup>H</sup> | 0.0004 | 0.14 | 0.35 | 0.06 | 0.32 |

<sup>H</sup>Adjusted for hypertension medication use at Reunion
<sup>G</sup>Adjusted for glucose-affecting medication use at Reunion
<sup>L</sup>Adjusted for lipid medication use at Reunion
Time refers to the following assessment timepoints: baseline, post-intervention, and reunion
Bolded text indicates statistical significance (*p* < 0.05 for main effects and *p* < 0.10 for interaction effects)
BMI indicates body mass index; HDL-C, high-density lipoprotein cholesterol; LDL-C, low-density lipoprotein cholesterol; V̇O<sub>2</sub>, oxygen consumption

#### Body Composition

Favorable changes in body weight, fat mass, waist circumference, and BMI were observed over time, although total fat-free mass declined significantly (all *p* < 0.0001). Significant intervention group by time interactions were observed for body weight (*p* < 0.0001), total fat mass (*p* = 0.01) and BMI (*p* = 0.0002), indicating trajectories of change in these outcomes differed across intervention groups over the follow-up period. A significant group by time interaction was also observed for percent fat mass and percent fat-free mass (both *p* = 0.08). As expected, sex was significantly associated with percent fat mass, percent fat-free mass, total fat-free mass, and waist circumference (all *p* < 0.0001). As illustrated in **Figure 1**, a significant sex by time interaction for total fat mass (*p* = 0.007) indicated fat mass trajectories differed between men and women over time. Significant sex by time interactions were also observed for body weight (*p* = 0.01), BMI (*p* = 0.07), and percent fat mass and percent fat-free mass (both *p* = 0.06).

**Figure 1.**
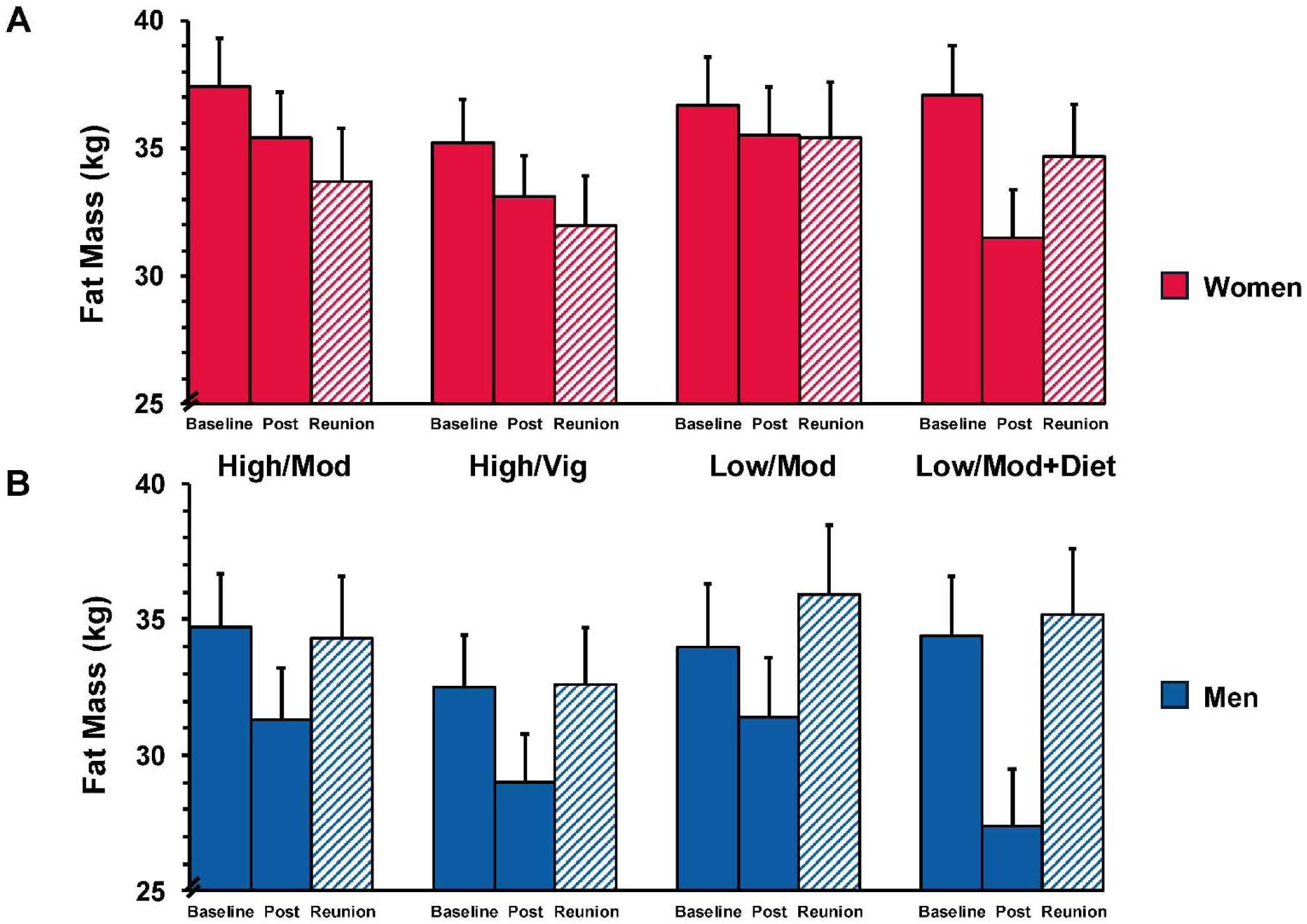
Least squares mean fat mass at baseline, post-intervention, and reunion by intervention group and sex. Bars represent least squares mean with standard error estimates from linear mixed effects models. Panel A: women; Panel B: men. A significant intervention group × time interaction (*p* = 0.01) and sex × time interaction (*p* = 0.007) were observed for total fat mass. High/Mod, high amount/moderate intensity aerobic training; High/Vig, high amount/vigorous intensity aerobic training; Low/Mod, low amount/moderate intensity aerobic training; Low/Mod+Diet, low amount/moderate intensity aerobic training plus diet-induced weight loss; Post, post-intervention.

#### Cardiorespiratory Fitness

Across all groups, both relative and absolute peak VϑO_2_ declined significantly over the follow-up period (both *p* < 0.0001). A significant main effect of intervention group was observed for relative peak VϑO_2_ (*p* = 0.04), indicating groups differed overall across the study period. For absolute peak VϑO_2_, a significant intervention group by time interaction was observed (*p* = 0.02), suggesting trajectories of decline differed across groups. Sex was significantly associated with both absolute and relative peak VϑO_2_ (both *p* < 0.0001), with men maintaining greater cardiorespiratory fitness across all timepoints. Significant sex by time interactions were observed for both absolute and relative peak VϑO_2_ (*p* = 0.002 and *p* = 0.05, respectively), indicating overall fitness trajectories differed between men and women over time. The divergence in absolute peak VϑO_2_ trajectories by sex is illustrated in **Figure S1**.

For descriptive comparison with previously reported findings from the STRRIDE I Reunion cohort,^13^ percent change in relative peak VϑO_2_ from baseline to Reunion is displayed by intervention group in **Figure 2**. All four groups demonstrated statistically significant declines from baseline (all *p* < 0.05), with mean percent changes of −16.4 ± 12.8% (High/Mod), −10.7 ± 11.7% (High/Vig), −10.4 ± 16.2% (Low/Mod), and −10.2 ± 16.9% (Low/Mod+Diet). Considerable within-group variability was observed, particularly in the low amount/moderate intensity-based groups.

**Figure 2.**
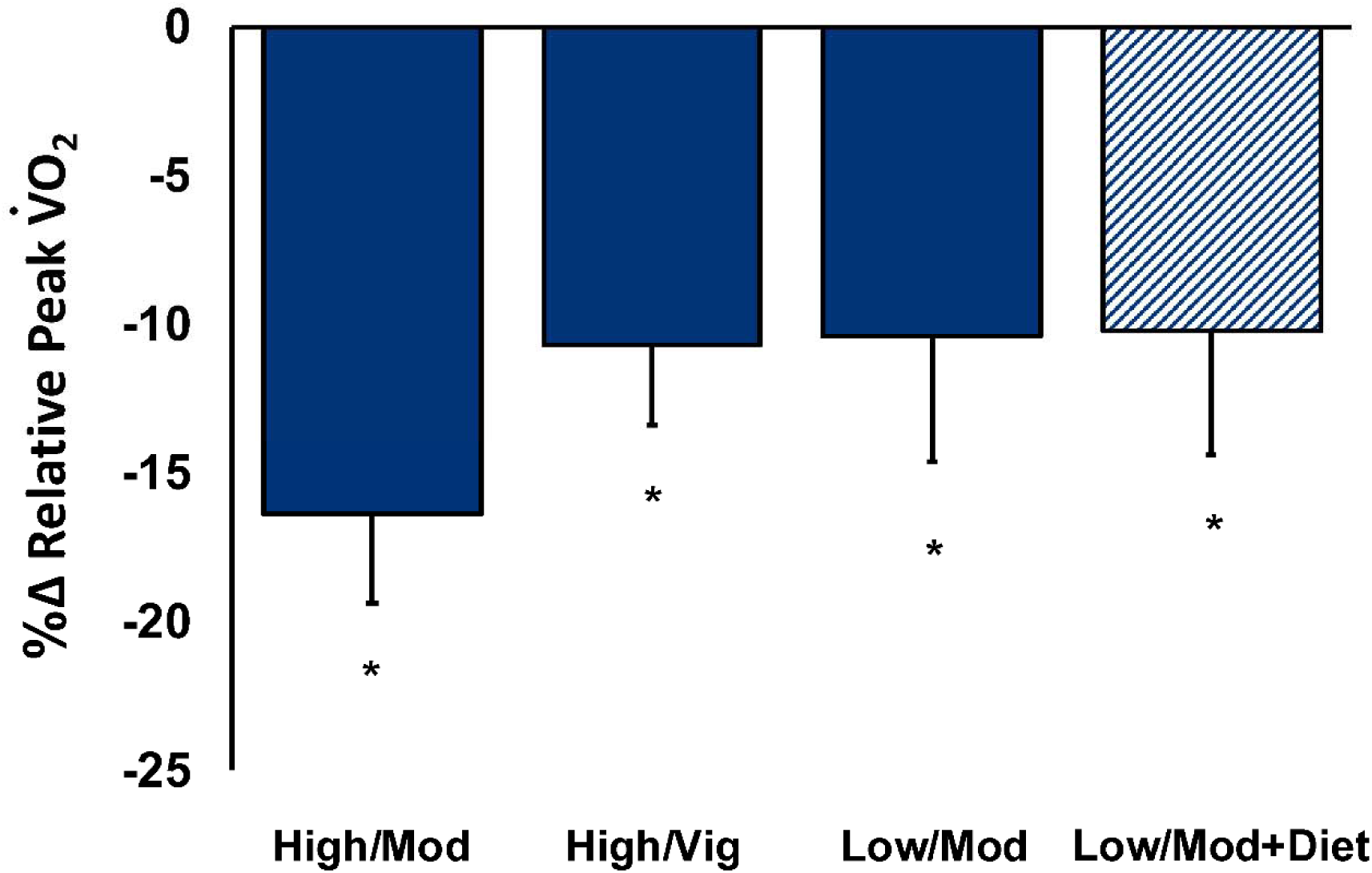
Percent change in relative peak VϑO_2_ from baseline to reunion by intervention group Bars represent mean percent change ± standard error. Percent change was calculated as [(Reunion value − baseline value) / baseline value] × 100. \**p* < 0.05, mean percent change significantly different from zero (one-sample t-test), indicating a significant decline in cardiorespiratory fitness from baseline to Reunion within each group. This analysis was performed for descriptive purposes to facilitate comparison with previously reported legacy effects on cardiorespiratory fitness in the STRRIDE I Reunion cohort.^13^ High/Mod indicates high amount/moderate intensity aerobic training; High/Vig, high amount/vigorous intensity aerobic training; Low/Mod, low amount/moderate intensity aerobic exercise; Low/Mod + Diet, Low/Mod plus diet-induced weight loss; VϑO_2_, oxygen consumption

To contextualize the clinical significance of observed fitness values, Reunion peak VϑO_2_ values were compared to previously described functional independence thresholds of approximately 18 mL/kg/min for men and 15 mL/kg/min for women.^27^ Among men, 3 of 25 Reunion participants (12%) fell below the functional independence threshold, with similar proportions across groups. Among women, the distributions appeared to differ by intervention group: no women in the High/Vig or Low/Mod+Diet groups fell below the 15 mL/kg/min threshold, compared with 1 of 11 women (9%) in the Low/Mod group and 3 of 10 women (30%) in the High/Mod group.

#### Cardiometabolic Blood-Based Markers and Hemodynamic Measures

HDL-C increased significantly over time (*p* < 0.0001). TG and total cholesterol also increased significantly (*p* = 0.001 and *p* = 0.03, respectively), while LDL-C remained stable over time (*p* = 0.97); models for LDL-C and total cholesterol were adjusted for lipid-lowering medication use at the Reunion timepoint. Sex was significantly associated with LDL-C (*p* = 0.001), total cholesterol (*p* = 0.001), HDL-C (*p* = 0.003), and TG (*p* = 0.03). Fasting glucose was stable over time (*p* = 0.10; adjusted for glucose-affecting medication use), although a significant sex by time interaction (*p* = 0.003) indicated glucose trajectories diverged between men and women over the follow-up period. A similar pattern was observed for TG (sex by time: *p* = 0.05).

Resting systolic and diastolic blood pressures increased significantly over time (both *p* = 0.0004); blood pressure models were adjusted for hypertension medication use. Mean arterial pressure differed significantly by sex (*p* = 0.04). MSSc decreased significantly over time (*p* < 0.0001; adjusted for hypertension and glucose-affecting medication use), indicating an overall improvement in composite cardiometabolic risk. A significant sex by time interaction for MSSc (*p* = 0.05) suggests trajectories differed between men and women over the follow-up period.

### Association Between Short-Term Intervention Response and Reunion Outcomes

To examine whether the magnitude of response to the original STRRIDE-PD intervention was associated with outcomes at the Reunion timepoint, linear regression models were fit for each outcome with and without adjustment for the corresponding baseline value (**Table 4**). In unadjusted models, short-term intervention response was not significantly associated with any Reunion outcome. However, after adjusting for baseline values, a markedly different pattern emerged, with significant positive associations observed across multiple outcome domains (**Table 4**).

**Table 4.** Unadjusted and Baseline-Adjusted Associations of Short-Term Intervention Response with Long-Term Reunion Outcomes.

| Outcome | Unadjusted |  | Adjusted for Baseline |  |
| --- | --- | --- | --- | --- |
|  | Coefficient (95% CI) | P-value | Coefficient (95% CI) | P-value |
| Body Weight (kg) | -0.89 (-1.86, 0.08) | 0.07 | 0.36 (-0.19, 0.90) | 0.20 |
| BMI (kg/m<sup>2</sup>) | -0.13 (-0.77, 0.51) | 0.69 | 0.28 (-0.23, 0.78) | 0.28 |
| Minimal Waist Circumference (cm) | 0.18 (-0.54, 0.90) | 0.62 | **0.70 (0.25, 1.16)** | **0.003** |
| Fat Mass (%) | 0.37 (-0.38, 1.12) | 0.33 | **0.56 (0.18, 0.94)** | **0.005** |
| Fat-free Mass (%) | 0.37 (-0.38, 1.12) | 0.33 | **0.56 (0.18, 0.94)** | **0.005** |
| Total Fat Mass (kg) | -0.21 (-0.79, 0.38) | 0.48 | 0.35 (-0.13, 0.84) | 0.15 |
| Total Fat-free Mass (kg) | 0.10 (-1.09, 1.29) | 0.86 | **0.84 (0.70, 0.99)** | **< 0.0001** |
| Absolute Peak V̇O<sub>2</sub> (L/min) | 0.16 (-0.44, 0.75) | 0.60 | **0.41 (0.10, 0.73)** | **0.01** |
| Relative Peak V̇O<sub>2</sub> (mL/kg/min) | 0.34 (-0.14, 0.82) | 0.16 | **0.40 (0.04, 0.75)** | **0.03** |
| Metabolic Syndrome z-score | -0.22 (-0.95, 0.50) | 0.54 | 0.02 (-0.48, 0.53) | 0.92 |
| Fasting Glucose (mg/dL) | 0.10 (-0.39, 0.58) | 0.69 | 0.31 (-0.16, 0.79) | 0.20 |
| HDL-C (mg/dL) | 0.24 (-0.74, 1.23) | 0.62 | **0.75 (0.24, 1.25)** | **0.004** |
| LDL-C (mg/dL) | -0.05 (-0.62, 0.52) | 0.86 | 0.38 (-0.43, 1.19) | 0.35 |
| Total Cholesterol (mg/dL) | 0.04 (-0.60, 0.68) | 0.90 | 0.46 (-0.37, 1.29) | 0.27 |
| Total Triglycerides (mg/dL) | 0.25 (-0.73, 1.23) | 0.61 | 0.67 (-0.38, 1.71) | 0.21 |
| Resting Mean Arterial Pressure (mmHg) | 0.03 (-0.32, 0.37) | 0.87 | 0.30 (-0.07, 0.66) | 0.11 |
| Resting Systolic Blood Pressure (mmHg) | -0.02 (-0.38, 0.34) | 0.91 | 0.12 (-0.25, 0.48) | 0.52 |
| Resting Diastolic Blood Pressure (mmHg) | 0.02 (-0.28, 0.33) | 0.88 | **0.47 (0.10, 0.84)** | **0.01** |

Bolded text indicates statistical significance ( $p < 0.05$ )
BMI indicates body mass index; HDL-C, high-density lipoprotein cholesterol; LDL-C, low-density lipoprotein cholesterol; V̇O<sub>2</sub>, oxygen consumption

For cardiorespiratory fitness, each 1 mL/kg/min greater improvement in relative peak VϑO_2_ during the intervention was associated with a 0.40 mL/kg/min higher relative peak VϑO_2_ at the Reunion timepoint among participants with equivalent baseline values (β = 0.40, 95% CI: 0.04-0.75; *p* = 0.03). A similar association was observed for absolute peak VϑO_2_ (β = 0.41 L/min, 95% CI: 0.10-0.73; *p* = 0.01).

After baseline adjustment among body composition and anthropometric outcomes, greater short-term improvements in percent fat mass (β = 0.56, 95% CI: 0.18-0.94; *p* = 0.005), total fat-free mass (β = 0.84, 95% CI: 0.70-0.99; *p* < 0.0001), and waist circumference (β = 0.70, 95% CI: 0.25-1.16; *p* = 0.003) were significantly associated with more favorable values at the Reunion timepoint.

After baseline adjustment among cardiometabolic blood-based markers, a greater short-term improvement in HDL-C was significantly associated with greater HDL-C at the Reunion timepoint (β = 0.75, 95% CI: 0.24-1.25; *p* = 0.004), while no significant associations were observed for TG, total cholesterol, LDL-C, or fasting glucose. After baseline adjustment for hemodynamic outcomes, greater short-term improvement in diastolic blood pressure was significantly associated with more favorable diastolic blood pressure at Reunion (β = 0.47, 95% CI: 0.10–0.84; *p* = 0.013), while associations for systolic blood pressure, mean arterial pressure, and BMI did not reach statistical significance.

## DISCUSSION

More than a decade after completing a 6-month exercise program, adults with prediabetes experienced lasting favorable effects on body composition and cardiometabolic health. With a unique framework to compare legacy effects across specific exercise doses and intensities, this study extends the emerging evidence base to a high-risk population at a critical inflection point for cardiovascular disease and type 2 diabetes progression. Illustrated by outcomes either improving or resisting expected age-related deterioration, these findings underscore the power and value of exercise training programs of even moderate duration for long-term health.

Body composition legacy effects were among the most striking findings of this study. First, waist circumference decreased from baseline to Reunion across all groups – suggesting durable attenuation of abdominal adiposity regardless of exercise dose, intensity, or diet-induced weight loss. Second, fat mass trajectories revealed compelling sex-specific patterns. Among men, total fat mass at the Reunion timepoint approximated pre-intervention baseline values regardless of randomization group – suggesting a potential attenuation of expected age-related fat mass accrual^28^ across all exercise conditions. Among women, a more compelling divergence emerged: those randomized to high amount exercise groups (High/Mod and High/Vig) continued reducing fat mass approximately 11 years after the intervention, while women in low amount groups, regardless of whether dietary intervention was included, showed trajectories more similar to men (**Figure 1**). Notably, the Low/Mod+Diet group showed an expected pattern of fat mass regain following initial weight loss, consistent with the well-documented challenges of long-term weight loss maintenance following combined lifestyle interventions.^12,29,30^ The mechanisms underlying this sex-specific response to high-volume exercise training on long-term fat mass trajectories remain unclear and represent an important hypothesis for future investigation, particularly given the potential implications for exercise prescription in women at elevated cardiometabolic risk.

Not all body composition findings told the same story: across all groups, fat-free mass declined significantly over the follow-up period. As a surrogate for skeletal muscle mass, the observed declines in fat-free mass are consistent with the progressive age-associated loss of skeletal muscle mass characteristic of sarcopenia – a condition associated with increased cardiovascular risk, functional decline, and mortality in older adults.^31^ Declines in fat-free mass may have contributed to the observed reductions in peak VϑO_2_ across groups, consistent with the well-established dependence of aerobic capacity on skeletal muscle mass in both sedentary and trained older adults.^32–34^ These findings underscore the importance of incorporating resistance and muscle-strengthening activities alongside aerobic exercise to preserve skeletal muscle function and attenuate age-related functional decline.

Surprisingly, as all groups experienced a significant cardiorespiratory fitness decline at follow-up (**Figure 2**), the protective effect of vigorous intensity training observed in the STRRIDE I Reunion^13^ was not replicated. Several factors may explain this discrepancy. The STRRIDE-PD cohort was older (71.3 vs. 63.0 years at Reunion) and living with prediabetes – a metabolic phenotype characterized by skeletal muscle insulin resistance associated with reduced aerobic capacity and attenuated exercise adaptability.^35,36^ Beyond age 70, the rate of cardiorespiratory fitness decline accelerates sharply.^37^ Thus, the cumulative effects of aging, prediabetes-related metabolic dysfunction, and fat-free mass loss over the follow-up period may have compounded fitness decline in ways not observed in the younger, lower-risk STRRIDE I cohort.

As cardiorespiratory fitness is one of the most powerful predictors of functional independence,^27^ cardiovascular disease, and mortality,^38,39^ the fitness declines observed in this cohort are clinically concerning — with consequences extending beyond cardiovascular risk to daily function and quality of life. At the Reunion timepoint, 7 of 68 participants had already fallen below cardiorespiratory fitness thresholds for functional independence,^27^ with others approaching these critical values. In this high-risk population, targeted exercise interventions – including re-engagement in structured training – represent an important strategy to restore and preserve functional independence.

Beyond group-level findings, exercise training may effectively reset an individual’s physiological baseline such that long-term health trajectories are shaped not by absolute change during the intervention alone, but by the new starting point from which aging-related decline proceeds. This concept is supported by a striking pattern in the intervention response analyses: while short-term intervention response was not significantly associated with any Reunion outcome in unadjusted models, after adjusting for baseline values, significant positive associations emerged across multiple health-related outcomes. In other words, two individuals achieving the same absolute improvement during the intervention may follow markedly different long-term health trajectories depending on where they started – revealing baseline health status as a critical determinant of long-term exercise benefits.

Consistent with findings from STRRIDE I, where greater short-term cardiorespiratory fitness gains were associated with greater fitness a decade later,^14^ the current study extends this observation to multiple health-related outcomes and to a population at considerably greater cardiometabolic risk. Significant baseline-adjusted associations spanned waist circumference, percent fat and fat-free mass, total fat-free mass, absolute and relative peak VϑO_2_, HDL-C, and diastolic blood pressure, reflecting the breadth of outcomes for which short-term improvements were associated with long-term benefit. Fat-free mass gains during the intervention were among the most durably preserved benefits over the follow-up period, whereby each 1 kg increase in fat-free mass during the intervention period was associated with 0.8 kg greater fat-free mass at the Reunion. These findings support a precision medicine approach to exercise prescription – identifying the right program, for the right person, for the right condition, at the right time – to maximize both short-term improvement and long-term cardiometabolic health.

### Limitations

Of the 175 original STRRIDE-PD completers, only 73 participated in the Reunion study, raising the possibility of selection bias. However, baseline characteristics and short-term intervention response measures did not differ meaningfully among Reunion participants and non-participants (**Table S1**), suggesting the findings are reasonably representative of the original trial completers. While the randomized design of the original trial supports causal inference for short-term intervention effects, the observational nature of the follow-up period limits causal inference; nevertheless, the intervention response analyses further support the exercise-induced legacy effect concept – short-term improvements appear to persist through physiological baseline resetting, direct physiological adaptation, and potential behavioral change. Physical activity throughout the follow-up period was not assessed; however, wearable-derived daily steps at the Reunion timepoint were positively associated with multiple cardiometabolic and fitness outcomes.^21^ Without a non-exercising control group, direct comparison of aging trajectories between previously trained and untrained individuals is not possible. As air displacement plethysmography does not disaggregate fat-free mass into specific tissue components, we could not directly determine changes in skeletal muscle mass. Body composition data were unavailable for the first five participants enrolled and six participants did not complete the cardiopulmonary exercise test, resulting in minor outcome-specific missingness. Given the exploratory nature of these analyses and small sex-specific group sizes, no correction for multiple comparisons was applied and findings should be interpreted with caution. Despite these limitations, the STRRIDE-PD Reunion study represents an important contribution to the emerging evidence base for exercise-induced legacy effects.

## Conclusion

More than a decade after completing a 6-month exercise program, adults with prediabetes experienced lasting legacy effects on body composition and cardiometabolic health. Considering an individual’s starting point, how much they improved during the program was associated with better long-term fitness, body composition, and cardiometabolic health – indicating exercise training may effectively reset physiological trajectories to confer durable cardiometabolic and fitness protection. Legacy effects differed between men and women, most notably for fat mass, where high amount exercise was associated with continued fat mass reductions in women over the follow-up period. These findings affirm structured exercise training, even of modest duration, is a powerful strategy to preserve cardiometabolic health and functional independence with aging.

## ACKNOWLEDGEMENTS

We would like to thank all of the STRRIDE-PD Reunion participants, staff members, and student interns.

## ETHICS STATEMENT

This study was carried out in accordance with the recommendations of the Duke University Institutional Review Board with written informed consent from all participants. All participants gave written informed consent in accordance with the Declaration of Helsinki. The protocol (Pro00014088) was approved by the Duke University Institutional Review Board.

## DATA AVAILABILITY

The raw data supporting the conclusions of this article will be made available by the authors without undue reservation. Requests to access the data should be directed to WEK.

## DECLARATION OF GENERATIVE AI IN THE MANUSCRIPT PREPARATION PROCESS

During the preparation of this work, the authors used Claude (Anthropic) to support language editing and content organization. After using this tool/service, the authors reviewed and edited the content as needed and take full responsibility for the content of the published article.

---

**Supplementary Figure S1.**
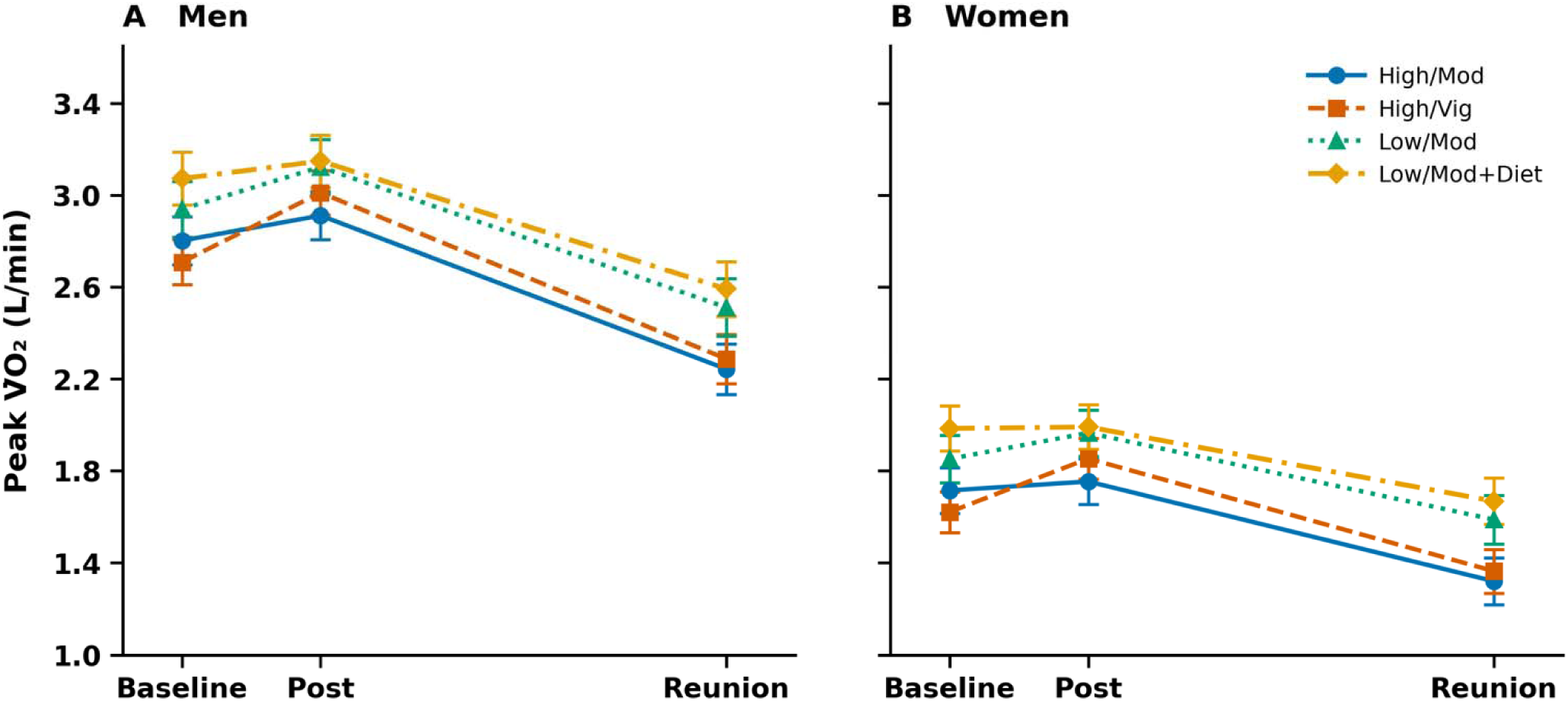
Absolute O_2_ at baseline, post-intervention, and reunion by intervention group and sex Values are least squares means ± standard error from linear mixed effects models. Significant intervention group × time and sex × time interactions were O_2_ (*p* = 0.02 and *p* = 0.002, respectively). The x-axis is not drawn to scale; baseline and post-intervention assessments occurred within a 6-month intervention period, while the Reunion assessment occurred approximately 11 years later. High/Mod, high amount/moderate intensity aerobic training; High/Vig, high amount/vigorous intensity aerobic training; Low/Mod, low amount/moderate intensity aerobic training; Low/Mod+Diet, low amount/moderate intensity aerobic training plus diet-induced weight loss; Post, post-intervention; O_2_, oxygen consumption.

**Supplementary Table S1.** Descriptive characteristics of the participants who completed the original STRRIDE-PD trial and those who completed the STRRIDE-PD Reunion study.

| Characteristic | STRRIDE-PD<br>Trial<br>(n=175) | STRRIDE-PD<br>Reunion<br>(n=73) |
| --- | --- | --- |
| **Intervention Group, n (%)** |  |  |
| High/Mod | 44 (25.1) | 18 (24.7) |
| High/Vig | 43 (24.6) | 23 (31.5) |
| Low/Mod | 43 (24.6) | 15 (20.5) |
| Low/Mod + Diet | 45 (25.7) | 17 (23.3) |
| **Demographics** |  |  |
| Age, mean (SD) | 59.2 (7.6) | 59.1 (7.2) |
| Women, n (%) | 108 (61.7) | 47 (64.4) |
| **Baseline Measures, mean (SD)** |  |  |
| Body Weight (kg) | 86.3 (12.4) | 86.0 (12.9) |
| BMI (kg/m<sup>2</sup>) | 30.4 (2.8) | 30.2 (2.8) |
| Minimal Waist Circumference (cm) | 98.6 (8.7) | 98.2 (9.4) |
| Fat Mass (%) | 41.0 (7.7) | 41.2 (7.6) |
| Fat-free Mass (%) | 59.0 (7.7) | 58.8 (7.6) |
| Total Fat Mass (kg) | 35.1 (7.4) | 35.4 (7.3) |
| Total Fat-free Mass (kg) | 51.3 (11.0) | 51.4 (11.3) |
| Absolute Peak V̇O<sub>2</sub> (L/min) | 2.15 (0.63) | 2.16 (0.65) |
| Relative Peak V̇O<sub>2</sub> (mL/kg/min) | 24.5 (5.2) | 24.7 (5.2) |
| Fasting Glucose (mg/dL) | 105.5 (9.8) | 104.4 (8.3) |
| HDL-C (mg/dL) | 46.9 (12.2) | 47.7 (11.0) |
| LDL-C (mg/dL) | 99.0 (20.5) | 98.7 (21.6) |
| Total Cholesterol (mg/dL) | 174.2 (27.2) | 173.0 (26.6) |
| Total Triglycerides (mg/dL) | 129.1 (70.7) | 119.2 (53.7) |
| Resting Systolic BP (mmHg) | 125.9 (13.4) | 123.6 (12.7) |
| Resting Diastolic BP (mmHg) | 75.2 (9.2) | 74.9 (9.8) |
| **Post-intervention Measures, mean (SD)** |  |  |
| Body Weight (kg) | 83.8 (11.8) | 83.3 (12.0) |
| BMI (kg/m<sup>2</sup>) | 29.5 (2.8) | 29.2 (2.8) |
| Minimal Waist Circumference (cm) | 97.1 (8.3) | 96.3 (8.9) |
| Fat Mass (%) | 38.9 (8.5) | 39.5 (7.9) |
| Fat-free Mass (%) | 60.9 (8.8) | 60.5 (7.9) |
| Total Fat Mass (kg) | 32.6 (8.0) | 32.8 (7.2) |
| Total Fat-free Mass (kg) | 51.5 (11.0) | 50.7 (11.1) |
| Absolute Peak V̇O<sub>2</sub> (L/min) | 2.27 (0.64) | 2.32 (0.67) |
| Relative Peak V̇O<sub>2</sub> (mL/kg/min) | 26.6 (5.5) | 27.4 (5.6) |
| Fasting Glucose (mg/dL) | 104.1 (9.8) | 103.8 (9.3) |
| HDL-C (mg/dL) | 48.3 (11.3) | 49.6 (11.3) |
| LDL-C (mg/dL) | 97.6 (20.6) | 98.1 (18.8) |
| Total Cholesterol (mg/dL) | 173.0 (28.5) | 173.2 (24.2) |
| Total Triglycerides (mg/dL) | 115.1 (55.1) | 109.5 (55.1) |
| Resting Systolic BP (mmHg) | 122.7 (14.0) | 121.4 (14.3) |
| Resting Diastolic BP (mmHg) | 73.0 (8.8) | 73.1 (8.8) |
| **Intervention Responses, mean change (SD)** |  |  |
| Body Weight (kg) | -2.7 (4.0) | -3.1 (3.6) |
| BMI (kg/m<sup>2</sup>) | -0.9 (1.4) | -1.1 (1.2) |
| Minimal Waist Circumference (cm) | -1.8 (4.1) | -2.7 (3.3) |
| Fat Mass (%) | -2.2 (3.7) | -2.4 (2.9) |
| Fat-free Mass (%) | 2.2 (3.7) | 2.4 (2.9) |
| Total Fat Mass (kg) | -2.9 (4.4) | -3.2 (3.7) |
| Total Fat-free Mass (kg) | 0.1 (2.3) | -0.1 (2.5) |
| Absolute Peak V̇O<sub>2</sub> (L/min) | 0.10 (0.20) | 0.13 (0.22) |
| Relative Peak V̇O<sub>2</sub> (mL/kg/min) | 2.0 (2.6) | 2.5 (2.5) |
| Fasting Glucose (mg/dL) | -1.4 (7.2) | -0.7 (6.0) |
| HDL-C (mg/dL) | 1.4 (5.4) | 2.0 (4.8) |
| LDL-C (mg/dL) | -1.3 (13.4) | -0.4 (12.6) |
| Total Cholesterol (mg/dL) | -1.0 (16.8) | 0.4 (14.4) |
| Total Triglycerides (mg/dL) | -13.8 (53.4) | -10.2 (34.9) |
| Resting Systolic BP (mmHg) | -3.5 (12.4) | -2.7 (11.2) |
| Resting Diastolic BP (mmHg) | -2.1 (8.1) | -1.6 (7.2) |

BMI indicates body mass index; BP, blood pressure; HDL-C, high-density lipoprotein cholesterol; High/Mod, high amount/moderate intensity aerobic training; High/Vig, high amount/vigorous intensity aerobic training; LDL-C, low-density lipoprotein cholesterol; Low/Mod, low amount/moderate intensity aerobic exercise; Low/Mod + Diet, Low/Mod plus diet-induced weight loss; V̇O<sub>2</sub>, oxygen consumption

